# Ranked (In)direct Citation Searching in Systematic Reviews: A methodological case study

**DOI:** 10.64898/2026.05.26.26354093

**Authors:** Tim Woelfle, Geoffrey Fucile, Julian Hirt, Rodrigo C. G. Pena, Magdalena Vogt, Thomas Nordhausen, Hannah Ewald, Christian Appenzeller-Herzog

## Abstract

Systematic Review (SR) is a prosperous study type in modern medicine and beyond. Many SR authors complement their primary database searches by supplementary techniques. Among these, citation-based techniques known as citation searching (CS) are widespread. Unranked Direct CS (UDCS) to identify directly cited and citing literature of seed references is currently most prevalent. Ranked (In)direct CS (RICS) additionally collects co-cited and co-citing literature combined with a ranking and cut-off procedure. However, RICS workflows remain non-standardized and tedious, and associated benefits unclear. This work aims to create a framework for the prospective international comparison of supplementary UDCS and RICS.

To prime RICS research, we developed the open-source *Co*Citation Network* application and assessed parallel supplementary UDCS and RICS retrospectively in three completed SRs and prospectively in one case study. Automated RICS collected and ranked cited, citing, co-cited, and co-citing literature of seed references from OpenAlex database and applied an empirical rank cut-off to approximate the volume of UDCS results. In RICS compared to UDCS, we consistently noted higher overlap with primary database search results. Title/abstract screening in the case study showed a precision (number needed to read) of 1.8% (57) for UDCS and 2.1% (48) for RICS results. After full text screening, two additional articles were included for review, one of which was identified by UDCS and RICS, and one exclusively by UDCS.

The present study indicates potential benefits of RICS for SR authors and will enable the formation of a research consortium to compare supplementary UDCS and RICS on larger scale.

**Highlights:** *What is already known?:*

- Citation searching is a widespread supplementary search technique in systematic reviews to complement the results of the primary literature search in bibliographic databases.
- Compared to traditional citation searching (UDCS), Ranked (In)direct Citation Searching (RICS) which additionally incorporates co-cited and co-citing literature with a relevance ranking and cut-off procedure has been proposed as a potentially more effective approach, but has only been evaluated in proof-of-concept studies.

*What is new?:*

- The open-source Co*Citation Network application is introduced: the first tool to automate parallel UDCS and RICS on the basis of the OpenAlex citation index.
- For the first time, parallel supplementary UDCS and RICS were applied and compared within a prospective systematic review, providing a concrete, replicable workflow.
- The results indicate that RICS results may be more relevant than UDCS results.

*Potential impact for RSM readers:*

- The proposed supplementary UDCS and RICS workflow and ready-to-use collaboration infrastructure allow systematic review teams worldwide to contribute data to a large-scale comparative study.

## 1. Introduction

### 1.1. Background and rationale

Upon framing their research question, most systematic reviewers initiate evidence retrieval by designing term-based literature searches in several bibliographic databases. These primary database searches are expected to be sensitive and of high quality, especially when they have been designed in collaboration with a librarian or information specialist.^1^ Nevertheless, adding one or several so-called supplementary search techniques to the primary database search can improve the comprehensiveness of evidence retrieval in a systematic review (SR).^2^ We herein use SR as umbrella term including all review types that are based on a systematic and transparent search methodology.^3^ Examples for supplementary search techniques include trial registry searching, web searching, hand-searching, contacting study authors or experts, and citation searching (CS).^4^

Standardization of CS workflows has recently been promoted by the publication of the *Terminology, Application, and Reporting of Citation Searching* (TARCiS) statement.^5^ The ten evidence-based TARCiS recommendations provide guidance on when and how to conduct CS and on how to report it. According to TARCiS, the seed references, i.e., the known articles that CS is based upon, should typically be identical to all eligible records that were derived from the primary database search. CS should then collect the directly cited and/or citing literature of these seed references, deduplicate this literature against the results of the primary database search (that have already been screened), and subject it to eligibility screening. The blunt volume of directly cited or citing literature retrieved by this standardized workflow is usually manageable, and we shall herein refer to this workflow as Unranked Direct CS (UDCS).

In addition to UDCS as the standard CS workflow, researchers have repeatedly proposed that collecting co-cited and co-citing literature of seed references may be a more effective method of identifying relevant publications for SRs.^6,7^ Co-cited literature (also known as co-citations) shares citing papers with a seed reference, co-citing literature (also known as bibliographic coupling) shares references with a seed reference (for illustration see ^5,8^). Along with their high potential relevance, these indirect citations composed of co-cited and/or co-citing papers are far more numerous than directly cited and citing papers,^6,7^ which hampers their seamless integration into standard SR workflows. In light of this, researchers have proposed different ranking methods and cut-off procedures to sort and prioritize direct and indirect CS results (cited, citing, co-cited, and co-citing papers) for evidence synthesis.^9,10^ We are collectively referring to these methods as Ranked (In)direct CS (RICS).

To our knowledge, application and evaluation of RICS for evidence retrieval in SRs has so far been restricted to proof-of-concept studies where published SRs were replicated by means of standalone RICS.^6,7,9,10^ CS is, however, commonly used as a supplementary search technique (see above) and current guidance explicitly discourages the use of standalone CS for SRs.^5^ Comparative studies on the effectiveness of UDCS and RICS that were conducted in a framework of supplementary SR searching are missing to date. Among the research priorities proposed by the TARCiS statement, research priority 1 reads: “The effectiveness, applicability, and conduct of indirect citation searching methods as supplementary search methods in systematic reviewing require further research (including retrieval of additional unique references, their relevance for the review and prioritization of results)”.^5^

Applications that automate CS rely on at least one citation index as the underlying database that provides information on citation linkages. Prominent citation indexes are Semantic Scholar,^11^ Lens.org, NIH iCite,^12^ OpenAlex,^13^ Web of Science (webofscience.com), or Scopus (scopus.com). While automated direct CS is widely available (e. g., citationchaser,^14^ Publish or Perish,^15^ or SpiderCite^16^), only few applications exist that automate indirect CS or RICS. These include the CoCites method and plugin that has been developed by the late Cecile Janssens on the basis of NIH iCite and is currently no longer being supported.^10^ Moreover, indirect citations of PubMed records can be automatically retrieved via the Citation Cloud that was built on the Anne O’Tate value-added search interface using NIH iCite data,^17,18^ the Connected Papers web application using Semantic Scholar data,^19^ the BibliZap application performing multi-level forward and backward citation searches with relevance-based ranking,^20^ and the Web of Science (via *“Related records”*) or Scopus (via *“Related documents based on references”*) database interfaces (the latter two retrieve only co-citing papers, not co-cited ones).

Our larger project “Ranked indirect versus unranked direct citation searching for evidence retrieval”^21^ is guided by the research question “*Does RICS display a higher effectiveness and efficiency than UDCS as a supplementary search technique in systematic reviewing?”*, with the key outcomes operationalized through a higher number of unknown relevant records (i. e., eligible records that have not been identified by the primary database search), a higher precision, and a lower number needed to read (NNR).^22^ Here, we describe the development of the Co*Citation Network and its utilization for parallel supplementary UDCS and RICS in a case study. The study workflow (Figure 1) pioneers the future work of a research consortium to generate conclusive evidence concerning our research question.

**Figure 1.**
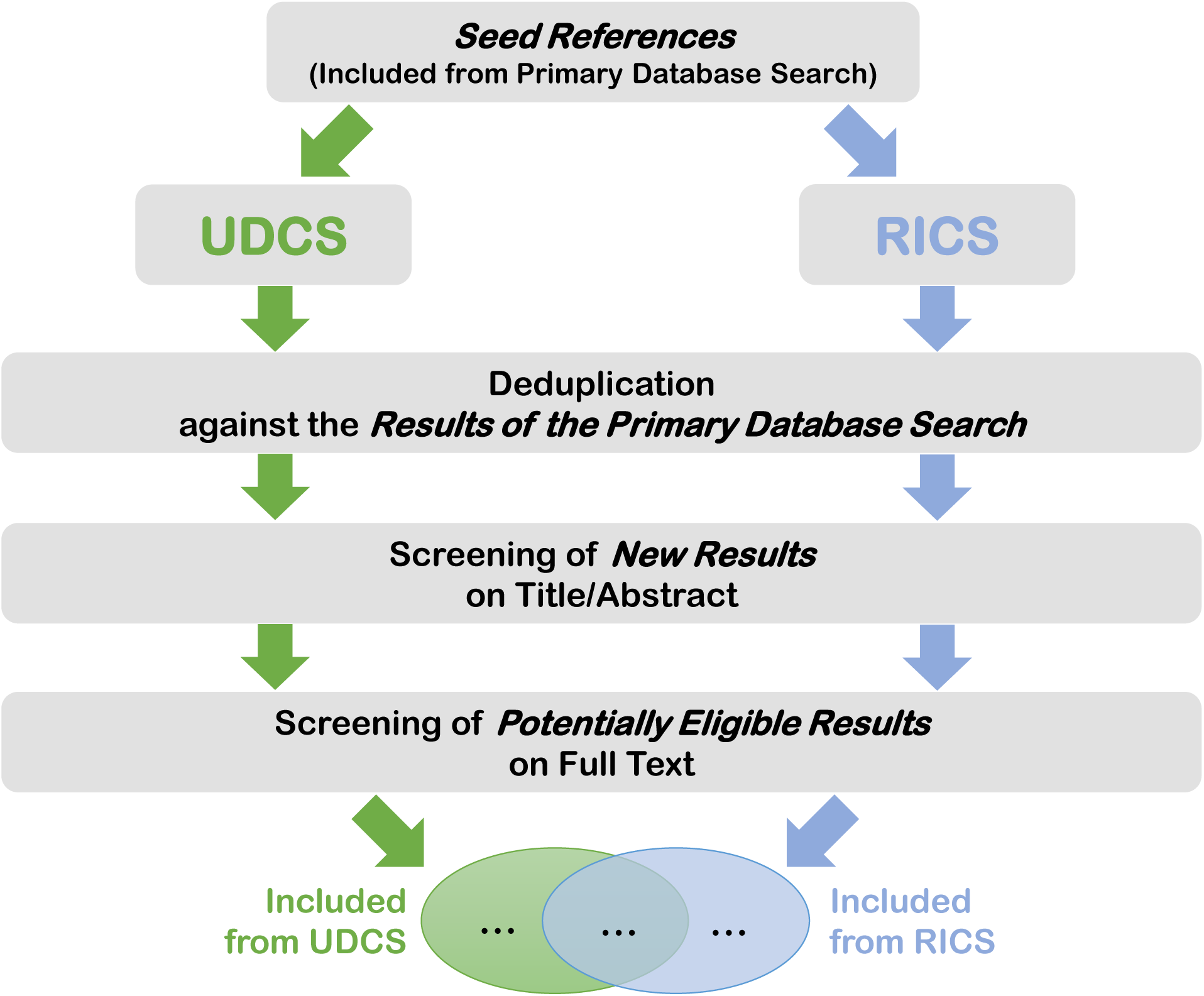
Workflow diagram of parallel supplementary UDCS and RICS in the framework of a systematic review project. RICS, Ranked (In)direct Citation Searching; UDCS, Unranked Direct Citation Searching.

### 1.2. Objectives

- To develop the Co*Citation Network as an application programming interface (API) including a flexible relevance rank cut-off that by default best matches the volume of directly cited and citing literature
- To exemplify a workflow of parallel supplementary UDCS and RICS, followed by eligibility screening of deduplicated results and prespecified evaluation of UDCS/RICS effectiveness and efficiency

## 2. Methods

### 2.1. Development of the Co*Citation Network application

To foster the exploitation of indirect citation relationships for evidence retrieval and SRs, we reasoned that the relative complexity of retrieval procedures could be mitigated by automation. Accordingly, the first prespecified step in this project was concerned with the development of an automation tool for RICS.^22^ Given that RICS collects the most relevant cited, citing, co-cited, and co-citing articles (see below), we termed this network tool Co*Citation Network to reflect its dual scope for direct and indirect citations.

The frontend of Co*Citation Network was integrated into the existing open source web application Local Citation Network.^23^ This application is an established tool for UDCS, providing both citing and cited records of the seed reference input directly from the OpenAlex application programming interface (API), among others.^13^ However, retrieving co-cited and co-citing records directly from the OpenAlex API is infeasible due to their much larger number compared to the direct citation links. We therefore implemented a local processing solution on our backend server: Snapshots of the OpenAlex academic graph are downloaded on a regular basis as data source. Identification and ranking/thresholding of cited, citing, co-cited, and co-citing references is performed locally and results are accessible to the frontend application via an internal API.

The source code of the web frontend of Co*Citation Network is open source (GPL-3.0 license) and available on https://github.com/LocalCitationNetwork/LocalCitationNetwork.github.io and https://github.com/CoCitationNetwork/CoCitationNetwork.github.io. The backend API service is developed in, and deployed from, a private repository. For transparency, however, we created a public release of the source code. This release is available on https://doi.org/10.5281/zenodo.20072168 (BSD 3-Clause License).

### 2.2. Relevance ranking method and rank cut-off

We used the relevance ranking method proposed by Belter.^9^ In this method, relevance ranks are calculated by summing up the number of citation relationships of a given publication with the seed reference(s). Four types of citation relationships – cited, citing, co-cited, and co-citing – are eligible, but since a publication hardly ever cites another publication and is cited itself by the same publication, a maximum of three citation relationships per seed reference can be assumed. Thus, a seed reference can cite a publication and concomitantly have both a co-cited and co-citing citation relationship with the same publication or it can be cited by a publication and have both a co-cited and co-citing citation relationship with the same publication. A supplementary RICS result based on three seed references can therefore achieve relevance ranks ranging from 1 to 9, and another search result based on ten seed references could have relevance ranks ranging from 1 to 30. A higher rank indicates a better match. For more details on the relevance ranking method and its rationale, refer to Belter.^9^

To be adopted into SR workflows, RICS must not vastly exceed the standard UDCS procedure with regard to the number of results and screening load. For this reason, we aimed for a default RICS relevance rank cut-off that will return approximately as many new records as would be retrieved by UDCS. Co*Citation Network was coded to concomitantly record RICS results with all relevance rank cut-offs and UDCS results. For the output of RICS results, the rank cut-off value that generates the number of results closest to and greater than the UDCS result is automatically chosen. Co*Citation Network users will – in addition to UDCS and RICS results for download – be provided with their project-specific, empirically determined default RICS rank cut-off value (flexibly allowing other cut-off values if preferred by the user). Choosing the next greater UDCS-adjacent RICS result was justified by the relatively higher overlap between the RICS- and the primary database search result (see below), which will more substantially reduce the RICS compared to the UDCS screening load by deduplication.

### 2.3. Overlap of UDCS- or RICS-with the primary database search results

The TARCiS statement recommends deduplicating supplementary CS results against the results of the primary database search before eligibility screening.^5^ A major fraction of relevant references retrieved by CS will thereby get eliminated, because they had already been retrieved by prior term-based searches. The higher the precision (i.e., the proportion of relevant documents or “relevance density”) in the CS results, the higher the overlap between CS and primary database search results is likely going to be. Belter showed that the precision in RICS results significantly increased with higher relevance ranks.^9^ Accordingly, we expected a higher precision and extent of primary database search overlap within the above-cut-off relevance ranks and, generally, when comparing RICS with UDCS results.

We set out to test these premises by running parallel supplementary UDCS and RICS searches. To this end, we retrospectively identified three completed SR projects where we had saved the results of the primary database searches in our records: Neuhaus et al.,^24^ Baroutsou et al.,^25^ and Reid et al.^26^ When extracting seed references, we found that 9/12 included records from Neuhaus et al., 14/14 included records from Baroutsou et al., and 24/25 included records from Reid et al. were indexed in OpenAlex database. Using an OpenAlex snapshot from November 26, 2024, UDCS and RICS results were generated in Co*Citation Network using the OpenAlex-indexed included records from each project as seed references (https://bit.ly/4x8ryar, https://bit.ly/4v9MU55, https://bit.ly/43yXZRq). Automatic RICS rank cut-off values were 5,^24^ 8,^25^ and 16,^26^ respectively. UDCS and RICS results were separately deduplicated for each SR against the results of the respective primary database search. For deduplication, we used a method that is largely identical to the Bramer method^27^ and has been documented for UDCS in an online video tutorial.^28^ This method eliminates those records as duplicates that have already been retrieved in the primary database search.

### 2.4. Prospective case study

To implement an UDCS versus RICS comparison in the context of a full supplementary literature search in a prospective case study, we conducted an SR assessing randomized and non-randomized trials on non-pharmacological interventions to support persons with young-onset dementia and their caregivers.^29^ The primary searches in five major bibliographic databases returned 2388 references, eleven of which met the project’s inclusion criteria.^30–40^ Together with four topical SRs and two study protocols identified during screening,^41–46^ these records were used as seed references for parallel UDCS- and RICS-based citation retrieval in the Co*Citation Network using an OpenAlex snapshot from March 13, 2025 (https://bit.ly/4dJQhZD). After deduplication, we subjected the UDCS and RICS results to separate eligibility screening by two independent reviewers using Rayyan.^47^ The screening method was split into a title-abstract-based and a full-text-based stage that, as recommended by the TARCiS statement,^48^ corresponded to the screening method that was used for the results of the primary database search. The workflow of parallel supplementary UDCS and RICS is illustrated in a flow diagram in Figure 1.

### 2.5. Deviations from protocol

We recorded the following deviations from our study protocol:^22^

1. We originally planned to generate RICS results using the new Co*Citation Network and UDCS results using the Local Citation Network.^23^ Since the latter tool uses the latest OpenAlex data via API while the Co*Citation Network relies on discrete snapshots of OpenAlex, the workflow in our protocol would have created a varying time gap between RICS and UDCS results. This time gap would have introduced bias when comparing the effectiveness of the two methods. By having Co*Citation Network to concomitantly provide both UDCS and RICS results based on the same OpenAlex snapshot, we have now removed this source of bias.
2. In the study protocol, we planned to apply a constant formula that was based on the number of seed references to calculate the rank cut-off of RICS results. In contrast to this, Co*Citation Network was coded to generate a default rank cut-off that is determined empirically by taking into account the number of UDCS results and RICS results with all relevance rank cut-offs.
3. The current version of the Co*Citation Network backend is implemented as an internal API rather than as a publicly accessible API. This deviation from the original protocol was necessary as the project does not have access to a sustainable funding source for the infrastructure required to operate a robust public-facing service.

## 3. Results

### 3.1. Retrospective analysis: RICS results may be more similar to primary database search results than UDCS results

Supplementary CS results should be as similar as possible to the seed references because widespread similarity to the seed references increases the chance of harboring unknown eligible content. In an SR, the primary database search is designed to identify the seed references, i. e. the records that meet the SR’s eligibility criteria. Therefore, if supplementary CS results resemble the primary database search results most closely, they are likely to be similar to the seed references and to harbor unknown eligible records. At the same time, similarity between the search results will manifest in a relatively high degree of overlap which is removed by deduplication in our workflow (Figure 1).

As shown in Table 1, the overlap of RICS results with the primary database search results was greater than that of UDCS results in all retrospective SR projects. This is consistent with our expectation that RICS results may be more similar to primary database search results and have a greater precision than UDCS results. The removal of a higher number of duplicates (records that have already been screened for eligibility by the research teams) led to a lower number of final CS results for a tentative eligibility screening in RICS compared to UDCS in 2 out of 3 SR projects (Table 1, column 2).

**Table 1.**
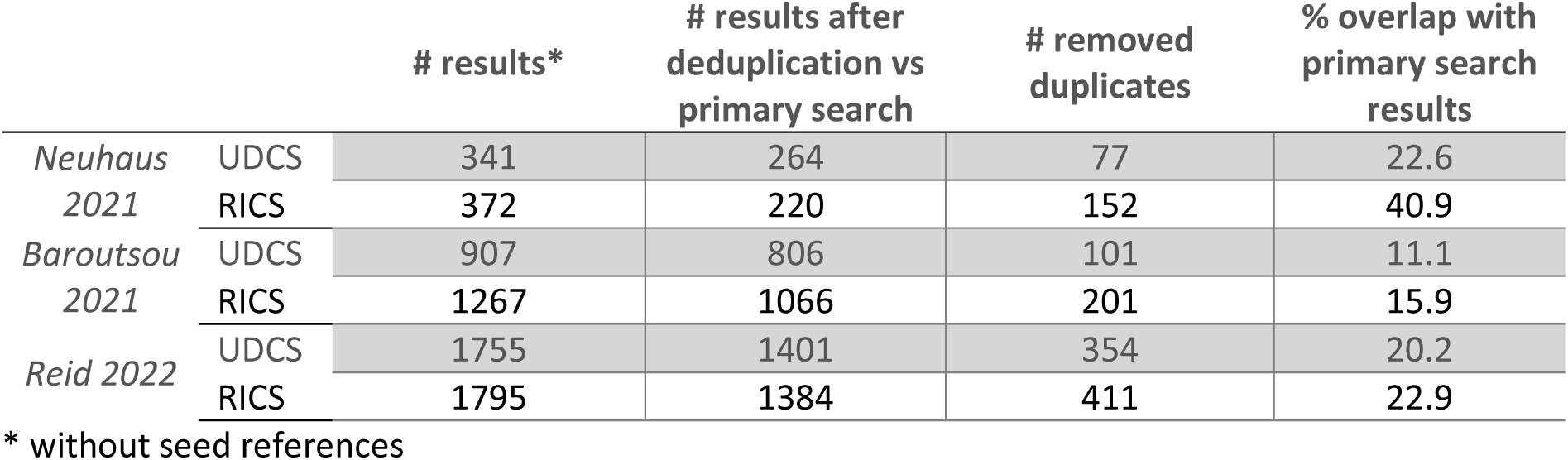

### 3.2. Prospective case study: Both UDCS and RICS identified additional eligible content

The results of supplementary CS are summarized in Figure 2. UDCS returned 917 results (corresponding to the number of unique records from 633 cited and 341 citing references), RICS – with an automatic rank cut-off of 10 – returned 1151 results (corresponding to the number of unique records from 283 cited, 114 citing, 786 co-cited, and 1147 co-citing references). Consistent with the retrospective analysis of three SRs (see above), the overlap with the primary database search was 7.5% and 12.9% for UDCS and RICS, respectively, leaving 848 and 1003 previously unseen UDCS and RICS records for eligibility screening.

**Figure 2.**
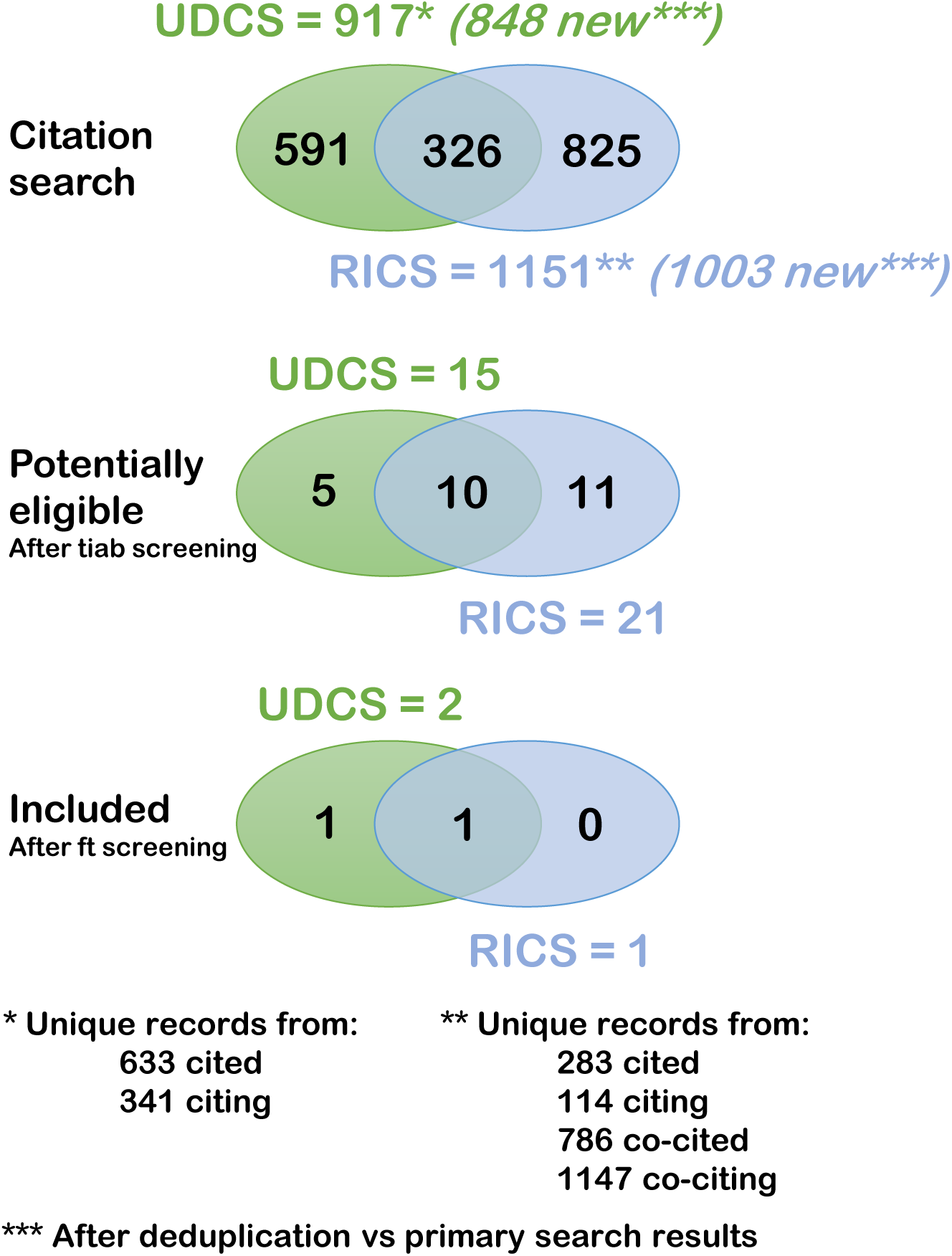
The volumes of UDCS (green) and RICS (blue) results in Vogt et al.^29^ are represented in Venn diagrams, illustrating their overlap after searching (citation search), after deduplication versus primary search results, after title/abstract screening (potentially eligible), and after full text screening (included). Note that UDCS results are composed of cited and citing references, and RICS results of cited, citing, co-cited, and co-citing references. Ft, full text; RICS, Ranked (In)direct Citation Searching; tiab, title/abstract; UDCS, Unranked Direct Citation Searching.

During title/abstract screening, 15 UDCS and 21 RICS records were deemed potentially eligible and were advanced to full text screening; ten of these records were identified by both methods. Full text screening identified one eligible record for inclusion that was identified by both methods^49^ and one additional eligible record that was exclusively identified by UDCS.^50^ Calculating from these figures, the precision of UDCS and RICS at the title/abstract screening stage was 1.8% and 2.1% which corresponded to a number needed to read (NNR) of 57 and 48, respectively. Precision and NNR with respect to final inclusion for review were 0.24% and 424 for UDCS and 0.1% and 1003 for RICS. After supplementary UDCS and RICS, the final SR included 13 records reporting on 9 studies.^29^

## 4. Discussion

To the best of our knowledge, RICS has never been applied in the context of supplementary SR searching.^51^ Publications that indicated the high potential of RICS to retrieve eligible literature with superior precision and varying recall effectiveness all have applied RICS as a stand-alone search technique.^6,7,9,10^ Thus, our explicit focus on supplementary CS in combination with a comparative UDCS/RICS workflow is novel. In addition to that, rigorous testing and comparison of different supplementary CS methods will likely feed into practice given the gold-standard recommendation that CS be only used as a supplementary method for evidence retrieval.^5^

So far, RICS research has been hampered by the lack of tailored automation tools. The automated RICS functionality of the Co*Citation Network now gives access to articles that rank high in the combined citation networks of any collection of seed references. Moreover, a purpose-built feature of the Co*Citation Network is the parallel performance of RICS and UDCS which will support our ongoing project of comparing supplementary UDCS and RICS on a broader scale.^22^ Thus, the Co*Citation Network forms the basis of a new workflow to evaluate the effectiveness and efficiency UDCS/RICS in the context of supplementary SR searching.

The present comparative case study does not have the power to clarify if UDCS or RICS shall be the method of choice for supplementary CS. Our objective was to exemplify a workflow of parallel supplementary UDCS and RICS evaluation as it can be embedded in other future SR projects (Figure 1). It is our hope that the adoption of this workflow by SR teams worldwide will enable a UDCS versus RICS comparison on a larger scale. For this reason, we have planned a prospective comparative study^22^ – the “RICS crowd study” – and deposited registration and results reporting forms with detailed instructions for collaboration on the Open Science Framework (https://osf.io/xutrn/overview). These can be completed and submitted by SR teams with the aim to be part of an international research consortium. We are confident that statistical evaluation of the consortium data will unveil whether or not RICS is more effective and efficient than UDCS for supplementary CS.

What can we learn on the relevance of UDCS and RICS results from the present case study? It is interesting and somewhat unexpected that one eligible study was exclusively identified by UDCS while the RICS technique did not yield any unique eligible records in this SR (Figure 2). This eligible study^50^ had a RICS rank of 6 (it was cited by one seed reference and co-cited by five other seed references) and, thus, clearly missed the automated rank cut-off of 10. Lowering the cut-off to 6 instead would have led to the retrieval of >10,000 RICS results, which we consider to be beyond acceptable specificity and feasibility. We conclude from this that RICS when tailored to the screening load of UDCS can miss relevant references that would have been captured using conventional UDCS, but we do not know if this is a frequent phenomenon. Whether or not this finding should end in the recommendation that both UDCS and RICS should routinely be used in parallel awaits the results of the prospective comparative study.

In the case study results, we also identify evidence to support our original premise that RICS results are more related to the seed references than UDCS results. Figure 2 shows that the overlap with the primary database search results was greater for RICS than for UDCS (148 versus 69 removed duplicates), as was the case for all retrospectively analyzed SRs (Table 1). Furthermore, RICS results harbored more unique potentially eligible records that passed the title/abstract screening stage than UDCS results (11 versus 5 records). Although none of the eleven unique RICS versus one of the five unique UDCS records finally fully complied with our SR eligibility criteria, these data may indicate a greater similarity of RICS over UDCS results with the seed references.

The way the Co*Citation Network performs the ranking and cut-off procedure according to Belter^9^ is only one of many possible ways to achieve this. Another citation-based possibility is the CoCites method.^10^ Further options include machine-learning- or large language model-based approaches that operate on the semantics of the seed references’ titles and abstracts.^52,53^ It is not the objective of this study to assess different possibilities of ranking and cutting-off indirect CS search results. Our principal rationale in favor of the Belter-procedure was to prioritize CS results with a method that is entirely based on citation relationships. As such, it maintains full complementarity to the primary database search by not mixing CS with a text-based approach that was already used for the primary search.

Using only one citation index, OpenAlex, as data source for the Co*Citation Network is a limitation, as different citation indexes always complement each other by filling at least some of the gaps that a single citation index inevitably creates.^5^ Pre-tests have shown that the citation index Lens.org via citationchaser^14^ consistently retrieved more cited and citing records of seed references than did OpenAlex (our unpublished data), which is consistent with current research.^54^ Furthermore, a deterioration of abstract coverage in OpenAlex has recently been reported.^55^ OpenAlex is, however, known as a rapidly evolving source that is on par with the established commercial citation indexes Web of Science and Scopus.^56^ Our choice to use OpenAlex was based on its methodological transparency and the free availability of periodic database snapshot downloads for academic use. Addition of further citation indexes as data sources is a viable option for future improvement should RICS indeed prove to be more effective than conventional UDCS for supplementary SR searching. In such a case, development of a public tool for automated multi-database RICS, which fully adheres to the TARCiS recommendations,^5^ is warranted.

## 5. Conclusions

This work provides the framework for a standardized use of RICS in SRs. Although preliminary, the results of our case study are consistent with the notion that RICS results may be more relevant, i. e. more related to the seed references, than UDCS results. The planned comparative study will, for the first time, address the question whether RICS is a suitable method to be included into the standard workflow of an SR project because it performs better as a supplementary search technique than UDCS.

## Data Availability

All data produced are available online

https://github.com/LocalCitationNetwork/LocalCitationNetwork.github.io

https://github.com/CoCitationNetwork/CoCitationNetwork.github.io

https://doi.org/10.5281/zenodo.20072168

https://osf.io/xutrn/overview

## Acknowledgements

Provision of resources and support for storing OpenAlex databases and serving the backend API service by sciCORE, the center for scientific computing at the University of Basel, is gratefully acknowledged.

## Declarations

### Ethics approval and consent to participate

Not applicable.

### Consent for publication

Not applicable.

### Availability of data and materials

The code of the web frontend of Co*Citation Network is open source (GPL-3.0 license) on https://github.com/LocalCitationNetwork/LocalCitationNetwork.github.io and https://github.com/CoCitationNetwork/CoCitationNetwork.github.io The code of the backend API service is openly deposited on https://doi.org/10.5281/zenodo.20072168 (BSD 3-Clause License). JSON files of all citation analyses were deposited on https://osf.io/xutrn/overview.

## Authors’ contributions

Conceptualization: JH, TN, HE, CAH

Data Curation: TW, GF, RCGP

Formal Analysis: JH, MV, CAH

Funding Acquisition: –

Investigation: JH, CAH

Methodology: TW, GF, JH, TN, HE, CAH

Project Administration: CAH

Resources: GF, RCGP

Software: TW, GF, RCGP

Supervision: JH, CAH

Validation: JH, CAH

Visualization: CAH

Writing – original draft: CAH

Writing – review & editing: all authors

